# Antenatal care attendance, intermittent preventive treatment in pregnancy and knowledge on malaria: A cross sectional study in a government and a private district Hospitals

**DOI:** 10.1101/2023.05.03.23289465

**Authors:** Charity Ahiabor, David Courtin, William Anyan, Atikatou Mama, Naa Adjeley Frempong, Kwadwo A. Kusi, Michael F. Ofori, Bright Adu, Bernard W. Lawson, Abraham K. Anang, Nicaise T. Ndam

**Author notes:** **Corresponding authors:** (CA), (NTN). Contribution.

## Abstract

Ghana has made significant strides in maternal healthcare under the National Health Insurance Scheme, but more effort is required to achieve the Sustainable Development Goal 3.1 target of <70 deaths/100,000 live births from the current 308 deaths/100,000. This study sought to investigate knowledge about ANC, ANC attendance, knowledge about malaria and IPTp among pregnant women, visiting a government Hospital in Mafi-Adidome (a rural community) and a private Hospital in Battor-Dugame (a rural-urban community) in the Volta region (a high malaria endemic area in Ghana).

A total of 1295 consenting pregnant women participated in the study. Information on sociodemographic characteristics, knowledge about ANC, IPTp and knowledge about malaria were collected by questionnaire. Chi-square tests were used to compare proportions of variables across the two study sites.

Higher proportion (70.8%) of participants accessing Battor Catholic Hospital showed adequate knowledge about ANC than 38.1% in Adidome Government Hospital (*X*^*2*^(7) =105.11; p<0.001). Also, higher numbers (46.6%) in Battor Catholic Hospital showed adequate knowledge on the timing for IPTp administration than 21.1% in Adidome Government Hospital (*X*^*2*^(2) =83.37; p<0.001). Although knowledge about malaria was generally low (0–33.3%) at both health centres, high proportions of participants (>80%) possess and use mosquito bed nets for malaria prevention. A lower proportion (46.6%) of participants in Battor Catholic Hospital made the WHO recommended 4-7 visits compared to 50.2% participants in Adidome Government Hospital. Age, marital status, employment and education influenced utilization of antenatal and delivery services. More sensitization on the importance of ANC and hospital delivery is needed in the study areas.

## Introduction

Maternal morbidity and pregnancy-related complications are a major health concern in developing countries. According to World Health Organisation (WHO) estimations, approximately eight hundred (800) women die each day from pregnancy and childbirth complications (1). Even though there is evidence of effective interventions which are available at reasonable cost for the prevention and treatment of life-threatening maternal complications, many women continue to die through pregnancy and childbirth-related complications, especially in rural areas (2) due generally to delays in seeking healthcare.

In 2003, the Government of Ghana introduced the Ghana National Health Insurance Scheme (NHIS); a social intervention program intended to provide equitable access and financial coverage for basic health care services (3,4). A maternal health policy implemented under the NHIS permits free registration for pregnant women and free access to antenatal care (ANC) and delivery service as well as three months post-delivery care. To be registered onto the NHIS, a pregnant woman will need to present proof of pregnancy such as current antenatal card, pregnancy test result endorsed by a doctor or midwife or an ultrasound scan report at her own cost.

Though Ghana has seen significant improvement in maternal healthcare under the NHIS (5,6), more effort is required to achieve the Sustainable Development Goal (SDG) 3.1 which proposes a maternal mortality ratio less than 70 maternal deaths per 100,000 live births by 2030. Delay in identifying pregnancy related complications has been reported as a major cause of maternal morbidity and mortality, especially in rural areas (7,8). Key challenges to addressing this issue include breaking barriers to accessing critical health services, addressing inadequate financial capabilities of families or mothers, proximity to the health facility, education and other sociocultural factors. According to Apanga *et al*. in 2018 (9),’a lack of medical and laboratory equipment and inadequate knowledge about the benefits of antenatal care are a setback to the provision of effective and quality maternal care in the Upper East region of Ghana’. In order to improve on the quality of antenatal care and to reduce the risk of still birth and other pregnancy complications the WHO also reviewed its initial recommended four antenatal visits to eight (1). These new guidelines seek to ensure healthy pregnancy period and effective transition to positive labour and childbirth. Regardless of these initiatives, some women still fail to complete the WHO recommended ANC visits (10), despite having good knowledge about ANC services.

Pregnant women in Ghana are expected to deliver in a hospital facility supervised by a skilled birth attendant, however, reports show low utilization of such services. According to Ganle *et al*. in 2019 (11), the factors influencing utilization include distance to the health facility, making at least four antenatal visits and registration for NHIS among others. Manyeh *et al*. in 2017 (12) also identified sociodemographic determinants of age, educational level, parity and economic status as some of the factors influencing delivery at Hospital facilities. There is limited data on how the local women in north and central Tongu perceive safe pregnancy and how they react to complications associated with pregnancy.

In areas of moderate to high transmission of malaria, the WHO recommended a package of interventions for control which include the promotion and use of insecticide treated nets (ITNs), the administration of intermittent preventive treatment with sulfadoxine-pyrimethamine (IPTp-SP) as part of antenatal care and appropriate case management through prompt and effective treatment of malaria in pregnancy.

Mafi-Adidome and Battor-Dugame are two district capitals in the Volta region that are endemic for malaria due to their proximity to the Volta Lake. Located within each district capital is a health facility namely Adidome Government Hospital (AGH); and Battor Catholic Hospital (BCH) which is privately owned and managed by the Accra Archdiocese of the Catholic Church under the office of the Metropolitan Archbishop of Accra. Even though these Hospitals are NHIS accredited, patronage for antenatal care and delivery services are generally low, particularly on non-market days when transport services are not readily available.

Ghana is a beneficiary of the ‘Roll Back Malaria’ program; a WHO initiative instituted as part of measures to reduce morbidity due to malaria by increasing the availability, coverage and usage of insecticide-treated mosquito nets (ITNs). There is limited information on the knowledge about malaria among the inhabitants and the use of such preventive measures in the study areas.

### Research question

What is effect of the knowledge of antenatal care (ANC) and malaria, intermittent preventive treatment in pregnancy (IPTp) and mosquito bed net (ITN) use among pregnant women at district Hospital in malaria endemic community to the attainment of SDG 3.1?

The **aim** of this study, therefore, was to investigate the ANC attendance, knowledge about ANC and knowledge about malaria and IPTp among pregnant women at two district Hospitals (government and private) in rural malaria endemic communities.

The objectives were to:

1. Evaluate knowledge of the importance of ANC, knowledge on malaria and IPTp among pregnant women visiting AGH and BCH.
2. Estimate ANC attendance, IPTp-SP doses administered and ITN use at the two health centres.

## Methods

### Study area and participant recruitment

Mafi-Adidome (Central Tongu District capital) and Battor-Dugame (North Tongu District capital) share a common boundary, hence, there is movement of people, goods and services between them. Battor-Dugame has a densely populated semi-urban structure with moderate social amenities compared to Mafi-Adidome which has scattered settlements with fewer amenities. The population of the Central Tongu District, according to the 2010 population and housing census, was 59,411 consisting of 53.2% females and 46.8% males. The general fertility rate is 94.9 births per 1000 women aged 15-49 years (13). The population of the North Tongu District was 89,777, consisting of 52.7% females and 46.3% males. The general fertility rate was 84.4 per 1000 women aged 15-49 (14).

Adidome Government Hospital (AGH) is the main referral centre within the Central Tongu District. The Hospital, though relatively small with limited modern healthcare infrastructure, offers general healthcare services. Its catchment area, however, extends well beyond the Mafi-Adidome Township and includes neighbouring towns and small communities such as Mepe, Volo, Mafi-Kumase, Bakpa, Battor etc. The Hospital encourages all patients to the facility to subscribe to the NHIS because of the general inability of the people to pay for the full cost of treatment. Other smaller privately owned clinics and CHPS (Community-Health and Planning Services) compounds are located within the district offering certain categories of healthcare.

Battor Catholic Hospital (BCH) is also the main referral centre in the North Tongu District. The Hospital offers specialised healthcare services and has facility for training Resident Doctors and House Officers. It is a much bigger Hospital compared to AGH in functionality and is also NHIS accredited. The immediate catchment population is about 41,437 inhabitants (14). BCH also receives patients from adjoining towns and communities such as Mepe, Osudoku, Ada East and West, Volo, Abutia, Mafi-Adidome etc. Patients to the facility are likely to pay a bit more for similar service in AGH. Other small clinics and CHPS compounds are sited within the North Tongu District.

### Ethics approval and consent to participate

Pregnant women visiting these two health facilities for antenatal care and for delivery from November 2016 to March 2019 were recruited after informed consent was obtained. The objectives of the study were explained to each participant and was presented with a written consent form for their signature or thumb print. An interpreter was engaged to explain the contents of the consent form to participants who could not read and write.

Ethical approval for the study was obtained from the Institutional Review Board of the Noguchi Memorial Institute for Medical Research (NMIMR-IRB CPN 071/15-16 amend. 2017) and the Ethical Review Committee of the Ghana Health Service (GHS-ERC 06/06/16).

### Data capture and Analysis

Information on age, gravidity status, residence, marital status, educational level, occupation and knowledge on malaria, prevention and risks during pregnancy were obtained using a structured questionnaire and analysed with STATA. Information on malaria infection status at ANC was obtained from participant’s own account during questionnaire interview. At delivery, malaria infection statuses were obtained from hospital records in participant’s ANC booklets. Total ANC attendance was also obtained from ANC booklets. For associations, the various variables/factors were compared with choice of facility using Chi-square tests. Where assumptions for Chi-square fail (frequencies <5), Fisher’s exact tests were used. Differences in means of continuous variables were tested between facilities using t-tests and where requirements for t-tests failed, particularly normality of the data, Wilcoxon Rank Sum tests were used. The p-value threshold for significance in the analysis was 0.05.

### Availability of data and materials

Data will be made available upon request.

### Funding

Institute of Research and Development (IRD)

## Results

A total of 1,295 consenting pregnant women, comprising 752 from AGH and 543 from BCH were recruited. Eight hundred and thirty-one (831) was during first ANC visit and 464 during delivery. Of the 831 ANC participants, 451 were from AGH and 380 from BCH while the 464 delivery participants consisted of 301 from AGH and 163 from BCH.

### Socio-demographic characteristics

The mean ages were 25.9(±6.5) years at AGH and 27.4(±6.0) years at BCH with the majority within the ranges of 20-29 and 30-39 years, respectively (Table 1). Chi-square test showed statistically significant differences in age across the health facilities (*X*^*2*^(3) =17.89; p<0.001). Whereas most pregnant women accessed ANC clinics close to their residence, a small proportion of them i.e., 1.1% from Adidome were captured accessing BCH for their first ANC while 7.8% from Battor were captured at Adidome. The study also found more pregnant women from ‘other communities’ accessing BCH (45.8%) than AGH (5.1%) for the first ANC visit. Majority of participants with basic education i.e. Primary and Junior High School accessed AGH than BCH, while majority of participants with secondary and tertiary education, accessed BCH than AGH (Table 1). Mean gestation at first ANC consultation were 14.6(±7.3) weeks at AGH and 16.4(±6.9) weeks at BCH (t=3.50, p<.001). A higher proportion of the multigravidae was observed at the two health centres.

**Table 1:** Socio-demographics characteristics of study participants

|  | Health facility, N (%) |  | Test statistic | P-value |
| --- | --- | --- | --- | --- |
|  | AGH | BCH |  |  |
| Age (years) | | | $X^2(3)=17.89$ | $<0.001$ |
| <20 | 91 (20.2) | 37 (9.7) |  |  |
| 20 – 29 | 228 (50.6) | 209 (55.0) |  |  |
| 30 – 39 | 124 (27.5) | 124 (32.6) |  |  |
| 40+ | 8 (1.8) | 10 (2.6) |  |  |
| Mean (SD) | 25.9(6.5) | 27.4(6.0) | t = -3.46 | <0.001 |
| Age at first pregnancy (years) |  |  | t = -3.52 | <0.001 |
| Mean (SD) | 20.8 (3.7) | 21.8 (4.1) |  |  |
| Residence | | | $\chi^2(2)=612.99$ | <0.001 |
| Adidome | 393 (87.1) | 4 (1.1) |  |  |
| Battor | 35 (7.8) | 202 (53.2) |  |  |
| Other | 23 (5.1) | 174 (45.8) |  |  |
| Marital status | | | $\chi^2(2)=83.92$ | <0.001 |
| Married | 301 (66.7) | 320 (84.2) |  |  |
| Single | 147 (32.6) | 34 (9.0) |  |  |
| Cohabiting | 3 (0.7) | 26 (6.8) |  |  |
| Highest formal education | | | $\chi^2(4)=37.96$ | <0.001 |
| None | 46 (10.2) | 43 (11.3) |  |  |
| Primary | 111 (24.6) | 78 (20.5) |  |  |
| JHS | 239 (53.0) | 165 (43.4) |  |  |
| SHS | 48 (10.6) | 53 (14.0) |  |  |
| Tertiary | 7 (1.6) | 41 (10.8) |  |  |
| No. of children | | | $\chi^2(5)=5.45$ | 0.364 |
| 0 | 154 (34.2) | 118 (31.1) |  |  |
| 1 | 112 (24.8) | 110 (29.0) |  |  |
| 2 | 73 (16.2) | 67 (17.6) |  |  |
| 3 | 50 (11.1) | 48 (12.6) |  |  |
| 4 | 35 (7.8) | 22 (5.8) |  |  |
| 5+ | 27 (6.0) | 15 (4.0) |  |  |
| Mean (SD)* | 1.6 (1.6) | 1.5 (1.5) |  |  |
| Gravida | | | $\chi^2(1)=1.95$ | 0.163 |
| Primigravidae | 141 (31.3) | 102 (26.8) |  |  |
| Multigravidae | 310 (68.7) | 278 (73.2) |  |  |
| Gestation at ANC (weeks) |  |  | t = 3.50 | <0.001 |
| Mean (SD) | 14.6 (7.3) | 16.4 (6.9) |  |  |
| Total | 451 (100.0) | 380 (100.0) |  |  |

\*Test on differences carried out on only the categorical version of the variable.
$\chi^2(df)$ = Chi-square (degrees of freedom), t = Student t-test.
JHS = Junior High School, SHS = Senior High School

Chi-square analysis showed a significant association between formal education and choice of health facility (*X*^*2*^(4) =37.96; p<0.001). More of the married participants compared to the single and cohabiting assessed ANC at BCH than AGH with a statistically significant difference observed in marital status and choice of health facility (*X*^*2*^(2) =83.92; p<0,001). Also, higher proportion of multigravidae reported at BCH while higher proportion of the primigravidae were observed at AGH.

### Accessibility of health facility

Even though a total of 65.2% of respondents did not consider the hospital far from their houses, only 12.4% of them got to their respective health facilities by foot (ie 19.5% at BCH and 6.4% at AGH; *X*^*2*^(9) =154.15; p<0.001). Those who reached the hospital by motorbike (Okada) constituted 50.1% (ie 63.6% at AGH and 34% at BCH). Only a small proportion accessed Hospital by boat/canoe (ie 6.9% at AGH and 1.8% at BCH). Travel time to Hospital showed majority of the study participants spent between 10 -29 minutes from home to the hospital. This comprised of 45% at AGH and 47.6% at BCH. Study participants who spend more than one hour to get to Hospital comprise 3.3% at AGH and 5.5% at BCH (Table 2). Concerning cost of transportation, majority of participants at the time of study spend between 1-5 cedis to get to the Hospital facility (ie 59.4% at AGH and 46.3% at BCH; *X*^2^(5) =78.52; p<0.001).

**Table 2:**
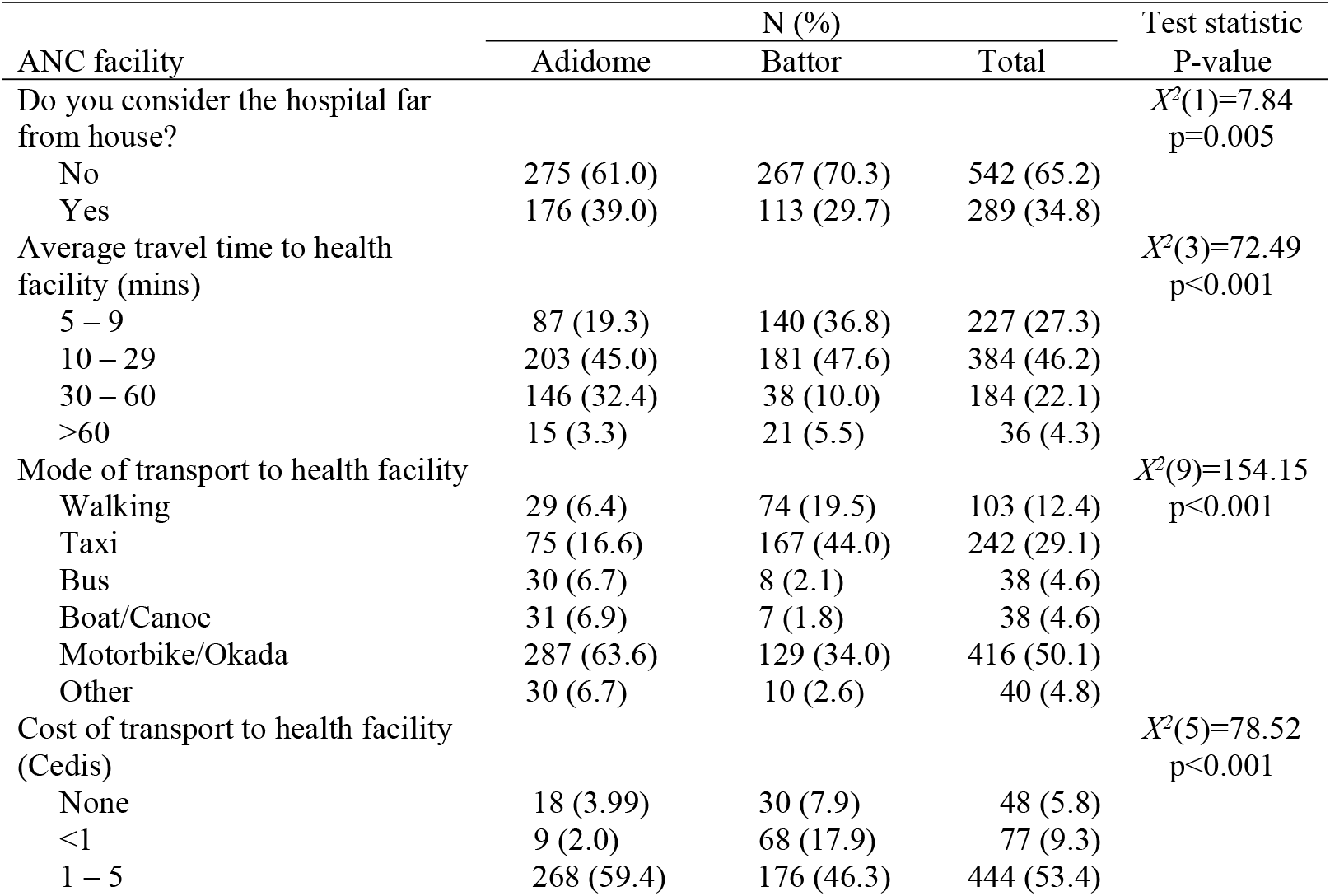
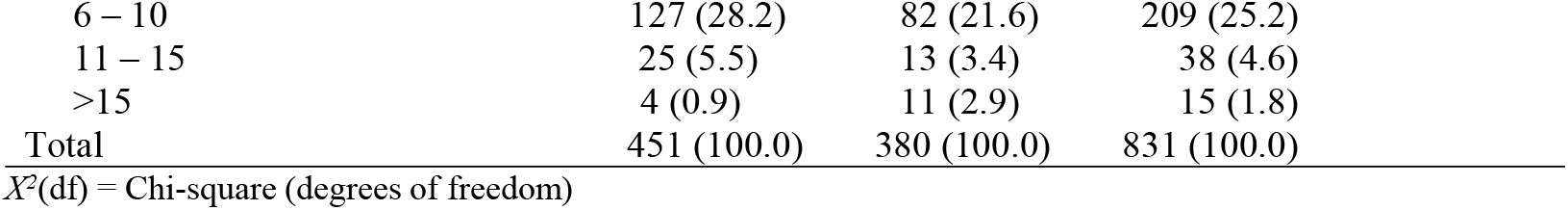
Accessibility of health facility

| ANC facility | N (%) |  |  | Test statistic<br>P-value |
| --- | --- | --- | --- | --- |
|  | Adidome | Battor | Total |  |
| Do you consider the hospital far from house? | | | | $X^2(1)=7.84$<br>$p=0.005$ |
| No | 275 (61.0) | 267 (70.3) | 542 (65.2) |  |
| Yes | 176 (39.0) | 113 (29.7) | 289 (34.8) |  |
| Average travel time to health facility (mins) | | | | $X^2(3)=72.49$<br>$p<0.001$ |
| 5 – 9 | 87 (19.3) | 140 (36.8) | 227 (27.3) |  |
| 10 – 29 | 203 (45.0) | 181 (47.6) | 384 (46.2) |  |
| 30 – 60 | 146 (32.4) | 38 (10.0) | 184 (22.1) |  |
| >60 | 15 (3.3) | 21 (5.5) | 36 (4.3) |  |
| Mode of transport to health facility | | | | $X^2(9)=154.15$<br>$p<0.001$ |
| Walking | 29 (6.4) | 74 (19.5) | 103 (12.4) |  |
| Taxi | 75 (16.6) | 167 (44.0) | 242 (29.1) |  |
| Bus | 30 (6.7) | 8 (2.1) | 38 (4.6) |  |
| Boat/Canoe | 31 (6.9) | 7 (1.8) | 38 (4.6) |  |
| Motorbike/Okada | 287 (63.6) | 129 (34.0) | 416 (50.1) |  |
| Other | 30 (6.7) | 10 (2.6) | 40 (4.8) |  |
| Cost of transport to health facility (Cedis) | | | | $X^2(5)=78.52$<br>$p<0.001$ |
| None | 18 (3.99) | 30 (7.9) | 48 (5.8) |  |
| <1 | 9 (2.0) | 68 (17.9) | 77 (9.3) |  |
| 1 – 5 | 268 (59.4) | 176 (46.3) | 444 (53.4) |  |

|  |  |  |  |
| --- | --- | --- | --- |
| 6 – 10 | 127 (28.2) | 82 (21.6) | 209 (25.2) |
| 11 – 15 | 25 (5.5) | 13 (3.4) | 38 (4.6) |
| >15 | 4 (0.9) | 11 (2.9) | 15 (1.8) |
| Total | 451 (100.0) | 380 (100.0) | 831 (100.0) |

$\chi^2(df) = \text{Chi-square (degrees of freedom)}$

### Knowledge about antenatal care and IPTp

During an assessment of participants’ knowledge on the importance of antenatal care, study participants were asked, ‘Why did you decide to visit the health centre?’ to which their responses include, ‘Feeling sick’, ‘Knowing the importance of antenatal care for the baby only’ and ‘Knowing the importance of antenatal care for the mother and for the baby’ (Table 3).

**Table 3:** Knowledge of importance of ANC and timing of IPTp

|  | ANC Health facility, N (%) |  | Test statistic | P-value |
| --- | --- | --- | --- | --- |
|  | AGH | BCH |  |  |
| Reason for attending ANC** | | | $\chi^2(7)=105.11$ | <0.001 |
| Feeling sick | 116 (25.7) | 66 (17.4) |  |  |
| ANC is important for mother | 93 (20.6) | 40 (10.5) |  |  |
| ANC is important for baby | 73 (16.2) | 9 (2.4) |  |  |
| ANC is important for mother & baby | 172 (38.1) | 269 (70.8) |  |  |
| Timing of IPTp during pregnancy | | | $\chi^2(2)=83.37$ | <0.001 |
| After first trimester exclusively | 95 (21.1) | 177 (46.6) |  |  |
| Anytime during the pregnancy | 2 (0.4) | 17 (4.5) |  |  |
| Don't know | 354 (78.5) | 186 (49.0) |  |  |
| Total | 451 (100.0) | 380 (100.0) |  |  |

\*\*Multiple responses allowed and so totals for variable may exceed totals below.
$\chi^2(df) = \text{Chi-square (degrees of freedom)}$ , IPTp = Intermittent Preventive Treatment in pregnancy

A statistically significant association was observed between ‘Reason for attending ANC’ and choice of health centre (*X*^*2*^(7) =105.11; p<0.001). Proportions of participants who responded ‘ANC was important for both mother and baby’ were 38.1% at AGH and 70.8% at BCH. The proportion of participants who responded ‘feeling sick’ were 25.7% at AGH compared to 17.4% at BCH. Similarly, when the question about the timing of IPTp administration was posed, 46.6% of ANC participants at BCH responded ‘after the first trimester exclusively’ as against 21.1% of ANC participants at AGH who responded same. Some study participants who responded, ‘don’t know’ were 78.5% at AGH compared to 49.0% at BCH (Table 3).

### ANC attendance and IPTp administration

Table 4 is a summary of information collated from the ANC booklets of participants at delivery. It includes information on total ANC visits made, doses of intermittent preventive treatment with sulfadoxine-pyrimethamine (IPTp-SP) administered, complications and emergency visits during the prenatal period. The number of ANC visits were categorized into participants who made less than four (<4) visits, between 4-7 visits and more than seven (>7) visits. Results show majority of participants at both health facilities made between 4-7 ANC visits with no statistically significant association observed between ANC attendance and health facility (*X*^*2*^(2) =1.17; p=0.558).

**Table 4:** ANC attendance and IPTp-SP doses administered.

| Delivery Health facility,<br>N (%) |
| --- |

|  | AGH | BCH | Test statistic | P-value |
| --- | --- | --- | --- | --- |
| Trimester at first ANC visit | | | $\chi^2(2)=1.67$ | 0.433 |
| 1st trimester | 136 (51.5) | 68 (45.0) |  |  |
| 2nd trimester | 108 (40.9) | 69 (45.7) |  |  |
| 3rd trimester | 20 (7.6) | 14 (9.3) |  |  |
| Total | 264 (100.0) | 151 (100.0) |  |  |
| Total no. of ANC visits | | | $\chi^2(2)=1.17$ | 0.558 |
| <4 | 70 (23.3) | 36 (22.1) |  |  |
| 4 –7 | 151 (50.2) | 76 (46.6) |  |  |
| 8+ | 80 (26.6) | 51 (31.3) |  |  |
| IPTp-SP given during current pregnancy | | | $\chi^2(1)=7.20$ | 0.007 |
| No | 27 (9.0) | 4 (2.5) |  |  |
| Yes | 274 (91.0) | 159 (97.6) |  |  |
| No. of IPTp-SP doses received | | | $\chi^2(4)=7.87$ | 0.096 |
| 1 | 43 (15.7) | 20 (12.6) |  |  |
| 2 | 49 (17.9) | 27 (17.0) |  |  |
| 3 | 111 (40.5) | 52 (32.7) |  |  |
| 4 | 48 (17.5) | 36 (22.6) |  |  |
| 5+ | 23 (8.4) | 24 (15.1) |  |  |
| Median (IQR) | 3.0 (2.0) | 3.0 (2.0) | $z=-2.17$ | 0.030<sup>r</sup> |
| When to take IPTp during pregnancy | | | $\chi^2(2)=11.97$ | 0.003 |
| After first trimester exclusively | 158 (52.5) | 83 (50.9) |  |  |
| Anytime during the pregnancy | 19 (6.3) | 26 (16.0) |  |  |
| Don't know | 124 (41.2) | 54 (33.1) |  |  |
| Emergency visits during current pregnancy | | | $\chi^2(1)=2.71$ | 0.099 |
| No | 271 (90.0) | 154 (94.5) |  |  |
| Yes | 30 (10.0) | 9 (5.5) |  |  |
| Complications during pregnancy | | | $\chi^2(1)=4.96$ | 0.026 |
| No | 278 (92.4) | 140 (85.9) |  |  |
| Yes | 23 (7.6) | 23 (14.1) |  |  |
| Total | 301 (100.0) | 163 (100.0) |  |  |

$\chi^2(df)$ = Chi-square (degrees of freedom), r = Wilcoxon Rank Sum test p-value

Findings on timing of first ANC visit showed 51.5% of participants reported at AGH and 45% reported at BCH in the first trimester (*X*^*2*^(2) =1.67; p=0.433). Only 7.6% in AGH and 9.3% in BCH reported in the third trimester. When the question about timing of IPTp administration was posed, higher proportions of study participants, such as 52.5% reporting at AGH and 50.9% reporting at BCH responded, ‘after the first trimester exclusively’ (*X*^*2*^(2) =11.97; 0.003). A lower proportion of participants ie 41.2% at AGH and 33.1% at BCH however, responded ‘don’t know’. Some study participants did not take IPTp-SP during the pregnancy period and these were 9% at AGH and 2.5% at BCH (Table 4). Only one person gave a reason for not receiving IPTp and that is, reporting late for the first ANC in the 38^th^ week of pregnancy. Very low percentages were recorded for complications and emergency visits at both health facilities (AGH and BCH) with p-values of 0.026 and 0.099 respectively across the health facilities.

### Knowledge about malaria, symptoms, risks and prevention

A score of participants’ knowledge about malaria, risks and prevention revealed a generally ‘low knowledge’ i.e., scores between 0-33.33% on the subject. Despite this observation however, a higher proportion of participants from BCH got ‘Medium’ and ‘High’ scores compared to participants from AGH. Among ANC participants, 55.2% from AGH and 21.3% from BCH scored ‘low knowledge’ (0-33.33%). A similar trend was observed at delivery ie 48.2% accessing AGH and 28.2% accessing BCH with a statistically significant corelation observed between knowledge about malaria and attendance at the health facilities both during ANC visits (*X*^*2*^(2) =124.37; p<0.001) and reporting for delivery (*X*^*2*^(2) =26.22; p<0.001).

Among ANC participants who scored ‘medium knowledge’ i.e. between 33.34–66.66%, a 69.2% was recorded at BCH compared to 44.8% at AGH. At delivery too, a higher proportion of 53.4% of ‘medium knowledge’ scorers was observed at BCH than at AGH (45.5%). High scoring (66.67-100%) study participants were only observed at BCH ANC and at the delivery wards of the two health centres, even though their numbers were low (Table 5).

**Table 5:** Level of knowledge on malaria causes, symptoms, risks, general prevention and prevention during pregnancy

|  | ANC Health facility, N (%) |  | Test statistic<br>P-value | Delivery Health facility, N (%) |  | Test statistic<br>P-value |
| --- | --- | --- | --- | --- | --- | --- |
|  | Adidome | Battor |  | Adidome | Battor |  |
| Knowledge level | | | $X^2(2) = 124.37$<br>$p < 0.001$ | | | $X^2(2) = 26.22$<br>$p < 0.001$ |
| Low | 246 (55.2) | 81 (21.3) |  | 145 (48.2) | 46 (28.2) |  |
| Medium | 202 (44.8) | 263 (69.2) |  | 137 (45.5) | 87 (53.4) |  |
| High | 0 (0.0) | 36 (9.5) |  | 19 (6.3) | 30 (18.4) |  |
| Total | 451 (100.0) | 380 (100.0) |  | 301 (100.0) | 163 (100.0) |  |
| Mean (SD) | 33.2 (9.8) | 46.6 (16.6) |  | 34.8 (20.0) | 48.5 (21.1) |  |

Knowledge level: Low = 0–33.33%, Medium = 33.34–66.66%, High = 66.67–100%

### Malaria and mosquito bed net usage

Information provided via responses to questionnaire and records from participant’s ANC booklets show a small proportion of participants both at ANC and at delivery had malaria during the study period. Among the ANC participants, 2.0% at AGH and 4.5% at BCH reported of malaria infection while 16.9% at AGH and 8.0% at BCH of delivery participants have records of malaria infection (Table 6). No confirmatory tests were however, carried out for 66.7% at AGH and 58.8% at BCH. Majority of participants possessed mosquito bed nets and a relatively high proportion of ANC participants i.e., 67.3% at AGH and 75.1% at BCH (p<0.001) compared to delivery participants ie 51.1% at AGH and 39.7% at BCH (p=0.039) used the nets regularly.

**Table 6:** Knowledge about Malaria and mosquito bed net usage among study participants

|  | ANC Health facility, N (%) |  | Test statistic<br>P-value | Delivery Health facility, N (%) |  | Test statistic<br>P-value |
| --- | --- | --- | --- | --- | --- | --- |
|  | Adidome | Battor |  | Adidome | Battor |  |
| Had malaria during current pregnancy | | | $\chi^2(2)=7.62$ | | | $\chi^2(1)=7.15$ |
| No | 425 (94.2) | 357 (94.0) | p=0.022 | 250 (83.1) | 150 (92.0) | p=0.007 |
| Yes | 9 (2.0) | 17 (4.5) |  | 51 (16.9) | 13 (8.0) |  |
| Don't know | 17 (3.8) | 6 (1.6) |  | NA | NA |  |
| *For those who had malaria,* |  |  |  |  |  |  |
| Malaria confirmatory test done |  |  |  |  |  |  |
| No | 6 (66.7) | 10 (58.8) |  | NA | NA |  |
| Yes | 3 (33.3) | 7 (41.2) |  |  |  |  |
| Have mosquito bed net | | | $\chi^2(1)=0.15$ | | | $\chi^2(1)=1.08$ |
| No | 29 (6.4) | 22 (5.8) | p=0.701 | 31 (10.3) | 12 (7.4) | p=0.298 |
| Yes | 422 (93.6) | 358 (94.2) |  | 270 (89.7) | 151 (92.6) |  |
| No. of bed nets | | | $\chi^2(4)=28.30$ | | | $\chi^2(4)=6.05$ |
| 1 | 225 (53.3) | 243 (67.9) | p<0.001 | 158 (58.5) | 93 (61.6) | p=0.167<sup>f</sup> |
| 2 | 140 (33.2) | 62 (17.3) |  | 70 (25.9) | 45 (29.8) |  |
| 3 | 46 (10.9) | 36 (10.1) |  | 29 (10.7) | 6 (4.0) |  |
| 4 | 5 (1.2) | 7 (2.0) |  | 7 (2.6) | 4 (2.7) |  |
| 5+ | 6 (1.4) | 10 (2.8) |  | 6 (2.2) | 3 (2.0) |  |
| Bed net use frequency* | | | $\chi^2(3)=16.05$ | | | $\chi^2(3)=7.97$ |
| Always | 284 (67.3) | 269 (75.1) | p<0.001<sup>f</sup> | 138 (51.1) | 60 (39.7) | p=0.039 |
| Sometimes | 131 (31.0) | 73 (20.4) |  | 123 (45.6) | 81 (53.6) |  |
| Seldom | 5 (1.2) | 8 (2.3) |  | 6 (2.2) | 9 (6.0) |  |
| Source of bed nets** | | | | | | $\chi^2(12)=28.49$ |
| ANC (hospital) |  |  |  | 258 (95.6) | 130 (86.1) | p=0.005 |
| Pharmacy |  |  |  | 3 (1.1) | 1 (0.7) |  |
| Market | NA | NA |  | 14 (5.2) | 5 (3.3) |  |
| Friend/Family |  |  |  | 4 (1.5) | 6 (4.0) |  |
| House distribution |  |  |  | 3 (1.1) | 12 (8.0) |  |
| Other sources |  |  |  | 17 (6.3) | 9 (6.0) |  |
| Slept under bed net previous night | | | $\chi^2(1)=1.93$ | | | $\chi^2(1)=3.37$ |
| No | 68 (15.1) | 71 (18.7) | p=0.165 | 132 (43.9) | 86 (52.8) | p=0.066 |
| Yes | 383 (84.9) | 309 (81.3) |  | 169 (56.2) | 77 (47.2) |  |
| Total | 451 (100.0) | 380 (100.0) |  | 301 (100.0) | 163 (100.0) |  |

\*Missing values accounted for but not shown. \*\*Multiple responses allowed and so totals for variable may exceed totals below.
NA = Not Applicable (was not part of questionnaire). $\chi^2(df)$ = Chi-square (degrees of freedom), f = Fisher's exact p-value.

## Discussion

Maternal age was observed to be a factor in ANC service utilization. Age at first ANC visit was mainly within 20–39 years and similar patterns were observed in the two health centres (Table 1). This follows the Ghana Health Service (GHS) recommendation which advocates 20 – 35 years as the peak reproductive years with minimum complications. This age category conforms to another study by Manyeh *et al*, (12) where the average age of a group of first time mothers in a rural community in southern Ghana was observed to be 23 years. Yaya *et al*.,(15) also reported the average age for ANC attendance in Ghana at 30.6 (±7.1) years.

Marital status was also observed to influence ANC service utilization, since high proportions of the married participants were observed at the two health facilities (Table 1). A similar finding was reported by Sakeah *et al*., (16) where marital status was associated with ANC attendance. Pregnant women who were single, widowed or divorced were less likely to make the recommended number of ANC visits (17). Pregnant women living in urban communities and supportive spouses in a marriage union may have a higher solvency and this may account for the higher numbers of married ANC participants observed in BCH where they were likely to pay a higher fee for a similar service in AGH.

The high proportion of Primary and Junior school leavers observed (Table 1) points to a high rate of school dropout in the two study areas. However, Battor-Dugame being a semi-urban area has more educational facilities compared to Mafi-Adidome hence the higher proportion of SHS and tertiary graduates observed at BCH. According to Muyunda *et al*., (18) women with higher education were more likely to make at least four ANC visits showing that education was associated with optimal ANC attendance. The high school dropout in the study areas, however, may account for the generally low ANC attendance. Even so, the higher numbers of basic school leavers observed in AGH may reflect a lower cost of ANC and delivery care offered at that facility.

Collectively, study participants at BCH appear better informed about the importance of ANC and timing of IPTp administration (Table 3). In spite of this observation, a lower proportion of 45.0% of BCH participants compared to 51.5% of participants at AGH initiated ANC visits in the first trimester in accordance with WHO recommendations. This observation can be compared to the finding of other studies in some rural communities in the southern and northern parts of Ghana (8,12) where only 50% of the study participants initiated ANC visits in the first and second trimesters, a trend which may have implications for maternal morbidity and mortality. Around 50% of the pregnant women made the recommended 4-7 ANC visits which is also comparable to the observation from other parts of Ghana (8,16,20).

Intermittent preventive treatment of malaria in pregnancy (IPTp) with sulfadoxine-pyrimethamine (SP) has been the WHO strategy for preventing malaria in pregnancy. This policy on IPTp-SP recommends uptake of at least three doses of IPTp-SP; each dose to be administered at a scheduled ANC visit. Results of this study show a median of three doses of IPTp-SP at each health facility (Table 4) showing compliance with the WHO recommendation of September 2012 for protection against malaria in pregnancy.

Pregnancy schools concept adopted by the Ghana Health Service has been an innovative and effective way to equip pregnant women with relevant knowledge pertaining to maternal and child health. In the study areas, these pregnancy schools are conducted during ANC visits with some success achieved especially in knowledge of timing of IPTp-SP among participants in AGH. At ANC enrolment, only 21.1% of study participants knew IPTp must be administered in the first trimester. However, at delivery the percentage of participants increased to 52.5% and this speaks to the continuous education given out at these pregnancy schools. The high percentage of participants who received IPTp-SP ie 91.0% in AGH and 97.6% in BCH is indicative of good coverage of IPTp administration at the two study sites.

Ownership and use of mosquito bed nets is one of WHO interventions towards reducing malaria transmission and morbidity within communities. Aside IPTp-SP, the high proportion of participants who possessed and used mosquito bed nets regularly pointed to the success of the Roll Back Malaria program in the study areas. The evidence of this could be due to the low prevalence of malaria and pregnancy related complications observed among the study participants at both study sites. The generally low level of knowledge about malaria observed was not seen to influence the prevalences observed.

## Conclusion

Higher proportion of participants below 20 years accessed Adidome Government Hospital while the older age groups of participants accessed Battor Catholic Hospital for obstetric care. Also, more married study participants were observed at Battor Catholic Hospital, whereas more of the single study participants were observed at Adidome. Majority of participants with basic (Primary, Junior High School) education accessed Adidome Government Hospital, whereas majority of participants with secondary and tertiary education, accessed Battor Catholic Hospital. Knowledge about the importance of ANC care was low especially in Adidome, however, knowledge about malaria was generally low in both study areas. Despite the NHIS intervention, attendance for delivery service was also generally low in both study sites, accessibility was observed to be a factor limiting attendance to Hospital. All study participants were NHIS subscribers, therefore, health insurance was not a factor limiting attendance. There is the need for more education on the significant impact of adequate ANC attendance on healthy pregnancy and use of Hospital delivery services to reduce maternal mortality in the study areas.

## Data Availability

Data will be made available on request

## Acknowledgements

Sincere thanks and gratitude to IRD (Institut de Recherche pour le Développement), funders of this study and to Mr. Tony Godi (School of Public Health, University of Ghana) for the analysis of the data. Our deepest appreciation to the staff of Battor Catholic Hospital and the staff of Adidome Government Hospital for all their assistance.

